# An individualized Bayesian method for estimating genomic variants of hypertension

**DOI:** 10.1101/2022.06.25.22276897

**Authors:** Md. Asad Rahman, Chunhui Cai, Dennis M. McNamara, Ying Ding, Gregory F. Cooper, Xinghua Lu, Jinling Liu

## Abstract

**Background:** Genomic variants of disease are often discovered nowadays through population-based genome-wide association studies (GWAS). Identifying genomic variations potentially underlying a phenotype, such as hypertension, in an individual is important for designing personalized treatment; however, population-level models, such as GWAS, may not capture all of the important, individualized factors well. In addition, GWAS typically requires a large sample size to detect association of low-frequency genomic variants with sufficient power. Here, we report an individualized Bayesian inference (IBI) algorithm for estimating the genomic variants that influence complex traits such as hypertension at the level of an individual (e.g., a patient). By modeling at the level of the individual, IBI seeks to find genomic variants observed in the individual’s genome that provide a strong explanation of the phenotype observed in this individual.

**Results:** We applied the IBI algorithm to the data from the Framingham Heart Study to explore genomic influences of hypertension. Among the top-ranking variants identified by IBI and GWAS, there is a significant number of shared variants (intersection); the unique variants identified only by IBI tend to have relatively lower minor allele frequency than those identified by GWAS. In addition, we observed that IBI discovered more individualized and diverse variants that explain the hypertension patients better than did GWAS. Furthermore, IBI found several well-known low-frequency variants as well as genes related to blood pressure that were missed by GWAS in the same cohort. Finally, IBI identified top-ranked variants that predicted hypertension better than did GWAS, according to the area under the ROC curve.

**Conclusions:** The results provide support for IBI as a promising approach for complementing GWAS especially in detecting low-frequency genomic variants as well as learning personalized genomic variants of clinical traits and disease, such as the complex trait of hypertension, to help advance precision medicine.

## Background

Hypertension (HTN) is a key risk factor for many cardiovascular diseases, and it was primarily responsible for about 7.8 million world-wide deaths in 2015 alone. Previous studies indicate that in addition to environmental factors, genomic factors play a significant role in blood pressure (BP) regulation [1]. Hypertension is a polygenic disease [2] burdening a large population across the globe. Current efforts at identifying significant genomic variants mostly involve the use of genome-wide association studies (GWAS). Although GWAS has successfully identified more than 1000 significant single nucleotide polymorphisms (SNPs; the most common type of genomic variants among people) for BP [3], there are limitations to this commonly used approach. In general, GWAS requires a large cohort to gain enough power to identify the significant SNPs, especially the ones with low minor allele frequency (MAF). That is why before 2015 there were only about 64 significant SNPs identified for blood pressure, and only recently were more SNPs identified due to the increased sample sizes (∼ 1 million individuals) [4-6]. Still, most of the SNPs identified so far are common SNPs with small effect sizes, and the total genetic variance in blood pressure explained by these ∼1000 SNPs is small (∼5.7%) [3]. It is likely that there are a significant number of non-common variants missed by GWAS that can help explain much of the remaining genomic variance [7].

GWAS is a population-based approach, and it extracts significant SNPs from a population level, not considering the specific genome of a given individual. Therefore, GWAS is not tailored to identify the genomic influences of HTN in an individual, which is the focus of personalized medicine. It is not uncommon that a HTN patient does not have any of the significant variants identified at the population level. Thus, identifying the most probable genomic variants of individual patients is important but remains an unmet need.

We have developed an individualized Bayesian inference (IBI) algorithm for estimating the genomic factors influencing the development of hypertension and other complex traits in a given individual. As a general machine learning framework, IBI applies a Bayesian method to identify the significant genomic variants in a given individual or patient. Bayesian methods including Bayesian multiple logistic [8] or linear regression have been used for identifying the causal SNPs among the significant genomic regions identified by GWAS (i.e., fine mapping) [9, 10]. However, none of these are individualized. IBI evolved from a tumor-specific causal inference algorithm (TCI) that members of our team developed for estimating the somatic mutations driving the development of individual cancerous tumors [11]. In contrast to TCI, IBI is designed to model and learn the relationships between an individual genome and a complex trait, such as HTN. Also, IBI was optimized for efficient computation with whole-genome data, whereas TCI was developed to use whole-exome data.

IBI identifies significant and potentially causal genomic variants for each individual based on his or her specific genomic background (and available training data on many similar individuals). By concentrating on the genomic variants observed in a particular individual, IBI has the potential to discover significant variants of low frequency that exist only in a small number of individuals and could have been missed by GWAS. The genomic variants identified being significant by IBI could help inform the design of personalized treatment for individuals with or at-risk for hypertension.

## Methods

### Overview of Bayesian Networks

A Bayesian network (BN) [12, 13] is a probabilistic graphical model which has two components. One is a graphical structure containing nodes and directed edges. Nodes represent domain variables such as genomic variants or clinical traits. Directed edges represent conditional dependencies between variables. The other component of a BN is a set of parameters which are conditional probabilities. Basically, each node has a conditional probability given its potential causes, which can be described by a conditional probability function. The joint probabilities of all nodes can be written as a product of each node’s conditional probability given its direct causes, based on the local causal Markov condition. A BN is a flexible framework for modeling the probabilistic relationships among variables in a complex domain via representing the joint probability of all the variables modeled in a probabilistic structure. A bipartite BN is a particular class of BN with less complexity, where there are only two sets of nodes in level 1 and level 2, and potential causal relationships only occur from nodes in level 1 to nodes in level 2.

How do we search for the most probable BN given data? A very popular class of methods are score-based algorithms that assign a Bayesian score to the BN model and return the BN with the highest score [12, 13]. This Bayesian score of the BN model is assigned based on how well this BN is supported by both the data and prior knowledge [14]. In this study, we will use a popular Bayesian score for modeling discrete variables, the Bayesian Dirichlet equivalent uniform (BDeu) score [14] as TCI did [11].

### The general framework of individualized Bayesian inference

As mentioned, IBI is based on TCI [11] and has been further developed and adapted to fit the circumstances of modeling a variety of complex diseases or traits and whole-genome genotyping or sequencing data. IBI is designed to estimate the significant genomic variants, such as SNPs, in a specific individual or patient for downstream clinical and molecular phenotypes. IBI uses a bipartite BN [12, 13] for modeling the conditional-dependency or predictive relationships between the genomic variants as a set of *V* nodes and the downstream traits or phenotypes as set of *T* nodes; directed edges between *V* and *T* nodes represent the probabilistic or predictive relationships from variants to traits (Figure 1A). Within this bipartite BN, among all the variants in one individual, IBI assigns a posterior probability for each variant (represented by an *V* node) in influencing or predicting the trait of interest (represented by node *T*) specific for this individual (Figure 1A, D).

**Figure 1.**
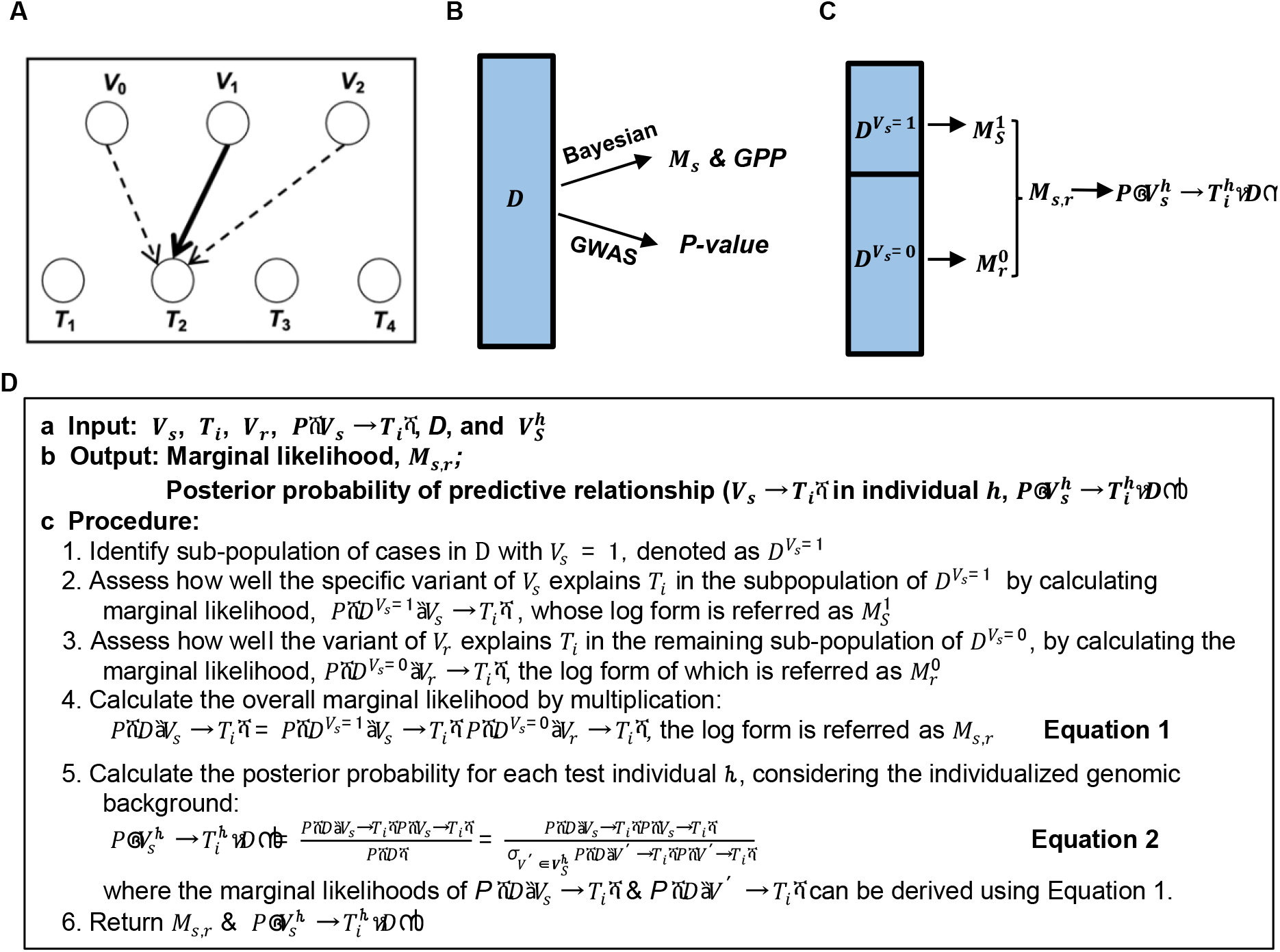
The IBI Algorithm. **A**. IBI uses a bipartite BN to model the probabilistic relationships from genomic Variants to Traits. *V* nodes denote variants and *T* nodes denote traits; the arcs denote the predictive relationships from *V* to *T* with only one assumed predictive variant being evaluated at a time indicated by the solid arc. **B**. Using the entire dataset (*D*) or population to evaluate the association between a particular genomic variant and a trait, GWAS methods output the p-value while the Bayesian method uses the marginal likelihood (*M*_*s*_) and global posterior probability (GPP). **C**. Based on the value of a particular variant *V*_*s*_, IBI partitions the whole population into two subpopulations, 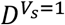 and 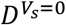, and derives the subpopulation-specific marginals, 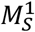 and 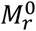, using *V*_*s*_ and *V*_*r*_ as the assumed cause specifically. The overall marginal *M*_*s,r*_ and the individual-specific posterior probability, 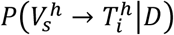 for the SNP *V*_*s*_ can be further derived. **D**. Pseudo code for the IBI algorithm.

For a current individual *h*, let *V*_*s*_ be a variable that represents a specific genomic variant *s* (e.g., a SNP) and let *T*_*i*_ be a specific trait *i* (e.g., HTN) of this individual. Let 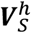 be a vector representing all the genomic variants in individual *h*. We will examine the predictive relationship (*V*_*s*_ → *T*_*i*_) in this individual for each possible *V*_*s*_. Let *P*(*V*_*s*_ → *T*_*i*_) be the prior probability for *V*_*s*_ predicting or influencing *T*_*i*_, which could be estimated using biological background knowledge or could be set using a uniform prior that assumes all the genomic variants have the same prior probability of predicting or influencing *T*_*i*_. Let *D* be the training data without inclusion of individual *h*. Let *M*_*s*_ represent the log form of the marginal likelihood of *P*(*D* |*V*_*s*_ → *T*_*i*_) that was derived by assuming one genomic variant *s* as the potential predictor for the entire population *D*; *M*_*s*_ can be further normalized by the summation of *M*_*s*′_ across all the SNPs to derive the posterior probability (PP). When scoring *V*_*s*_ → *T*_*i*_ for the entire population *D* (i.e., it is <u>not</u> individualized) using Bayesian learning and a uniform prior, PP is proportional to *M*_*s*_; thus, the ranking of the specific driver *V*_*s*_ by *M*_*s*_ or PP as a potential predictor of a trait at the population level is the same. Thus, we will use *M*_*s*_ as the score for *V*_*s*_ → *T*_*i*_. When evaluating the effect of *V*_*s*_ on *T*_*i*_ in the entire population using GWAS, the p-value is derived to indicate the significance of the association between *V*_*s*_ and *T*_*i*_ (Figure 1B).

IBI partitions the overall population into two subpopulations (Figure 1C). Suppose the current patient has *V*_*s*_ = 1, which represents the minor-allele of this SNP. Let 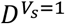 represent the patient-like-me subpopulation, where all the patients in this subpopulation contain the value *V*_*s*_ = 1. IBI evaluates how well *V*_*s*_ predicts the HTN status within 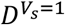, which has a marginal likelihood score of 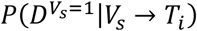 that we abbreviate as 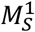 (Figure 1C, D). Let 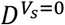 represent the remaining cases that do not have *V*_*s*_ = 1, but rather, have *V*_*s*_ = 0. To predict the data in 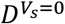, IBI finds the SNP *V*_*r*_ (where “r” denotes the remaining cases) that maximizes the marginal likelihood of *V*_*r*_ → *T*_*i*_, namely, 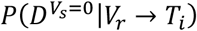, which we abbreviate as 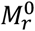. The marginal likelihood for all of the data, given *V*_s_ as an individualized predictor and *V*_*r*_ as the best predictor of the remaining cases, is 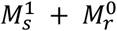, which we refer to as *M*_*s,r*_ (Figure 1C, D). This score of *M*_*s,r*_ can be used to evaluate and rank the capability of *V*_*s*_ in explaining the patients-like-me subpopulation that contain this minor allele as well as in helping reduce the noises for the remaining subpopulation.

The marginal likelihood is computed using the BDeu score [14] (Figure 1C, D; refer to the TCI paper [8]). Individualized posterior probabilities of the form *V*_*s*_ → *T*_*i*_ are further derived relative to the SNPs that are minor alleles in the genome of the current patient *h*. Thus, the posterior probability takes into consideration the specific genomic background of the given individual (Figure 1D, Equation 2). In summary, IBI is individualized in the following ways: (1) The overall marginal likelihood for each arc *V*_*s*_ → *T*_*i*_ (Equation 1) contains an individualized component that uses the subpopulation of “patients like me” that have the same variant (i.e., *V*_*s*_ = 1). (2) Each individual has a unique set of genomic variants. Depending on the specific set of variants, the posterior probability for the same arc of *V*_*s*_ → *T*_*i*_ may be different in different individuals (Equation 2). The individualized nature of IBI makes it a potential tool for advancing precision medicine where personalized treatments are desired for individuals of varying genetic backgrounds. IBI is implemented in python with vectorization and matrix operations for efficient computation involving millions of variants, and has been tested on whole genome sequencing data on the BioData Catalyst platform [15].

### Genome-Wide Association Studies

GWAS is the standard approach for identifying the significant variants associated with traits at the population level (e.g., p-value < 5 × 10^−8^ for genome wide significance). Conventional GWAS uses standard logistic regression models or Fisher’s exact test for discrete traits [16]. We performed GWAS using Fisher’s exact test on the same datasets to which we applied IBI and compared results.

### Data and data preprocessing

We used the whole genome genotyping data of Affymetrix HuGeneFocused50K from the Framingham Heart Study (FHS) cohort (dbGaP Study Accession: phs000007.v30.p11), which covered about 50K gene-centric and coding SNPs across the genome [17, 18]. We used the following functions from plink for further filtration and quality control, and have acquired 38,342 SNPs: --mind 0.03 --geno 0.03 --maf 0.01 –hwe 10e-6 –me 0.05 0.1 – sexCheck. We filled missing SNP values with the most frequent value for that particular SNP across the entire population. Dominant coding was then performed in plink, and thus, the final SNP values are 0 or 1 where 0 represents zero copy of the minor allele (risk allele) and 1 represents one or two copies of the minor allele. The focus of this paper is to predict the risk (minor) allele SNPs that influence hypertension (high blood pressure) rather than protecting the subject from hypertension. Therefore, we further removed the SNPs that have risk ratio smaller than 1 resulting in a total of 19,276 SNPs of interest.

Clinical phenotype data included harmonized systolic BP (SBP) and diastolic BP (DBP) data which were downloaded from PIC-SURE on the NHLBI BioData Catalyst platform [15]. SBP and DBP are specifically harmonized by the Trans-omics for Precision Medicine (TOPMed) Data Coordinating Center [19] by taking the average of two SBP or DBP measurements obtained at a single clinic visit. 10 and 5mm Hg were specifically added for SBP and DBP for individuals who were taking antihypertensive drugs [20]. If SBP>=140, or DBP>=90 or an individual who was taking antihypertensive drugs, we considered this individual as having HTN and assigned ‘HTN = 1’; otherwise, we classified this individual as not having HTN, and we assigned ‘HTN = 0’. After merging the SNP data and BP data, we obtained a total of 6,613 patients with 19,276 SNPs. We performed a stratified random split to produce an 80% training set (5,290 subjects) and a 20% test set (1,323 subjects), and we reserved this test set for the prediction task.

## Results

To evaluate IBI in inferring significant genomic variants for HTN, we compared its performance to that of GWAS. As a proof of concept, we applied both IBI and GWAS to the whole genome data of Affymetrix HuGeneFocused50K measurements and harmonized phenome data of BP measurements from the FHS cohort [18] as described above.

### A Bayesian method for GWAS analysis

As was explained in the Methods section, when using a Bayesian method and a uniform prior to study a single variant’s effect on HTN in the population level, the derived *M*_*s*_ for this variant is proportional to its global posterior probability (GPP), making it possible to use the marginal likelihood to find the top predicting SNP (Figure 2A). We observed that when using a uniform prior, as we did in this study, a population-based (i.e., not individualized) Bayesian approach to identifying top-ranked SNPs based on *M*_*s*_ yielded similar results to the population-based GWAS method (Figure 2A). The Spearman correlation coefficient between the p-value and *M*_*s*_ across all the 19,276 SNPs is -0.9. We further examined and compared the top 189 SNPs ranked by *M*_*s*_ or p-value. The top SNPs identified by high *M*_*s*_ values or low p-values are highly overlapping: 164 out of the top 189 SNPs and 18 out of the top 20 SNPs overlapped between these two rankings (Figure 2). Furthermore, the ranking of these top SNPs by *M*_*s*_ and p-value are either exactly the same or very similar where *M*_*s*_ is negatively correlated with p-value (Figure 2).

**Figure 2.**
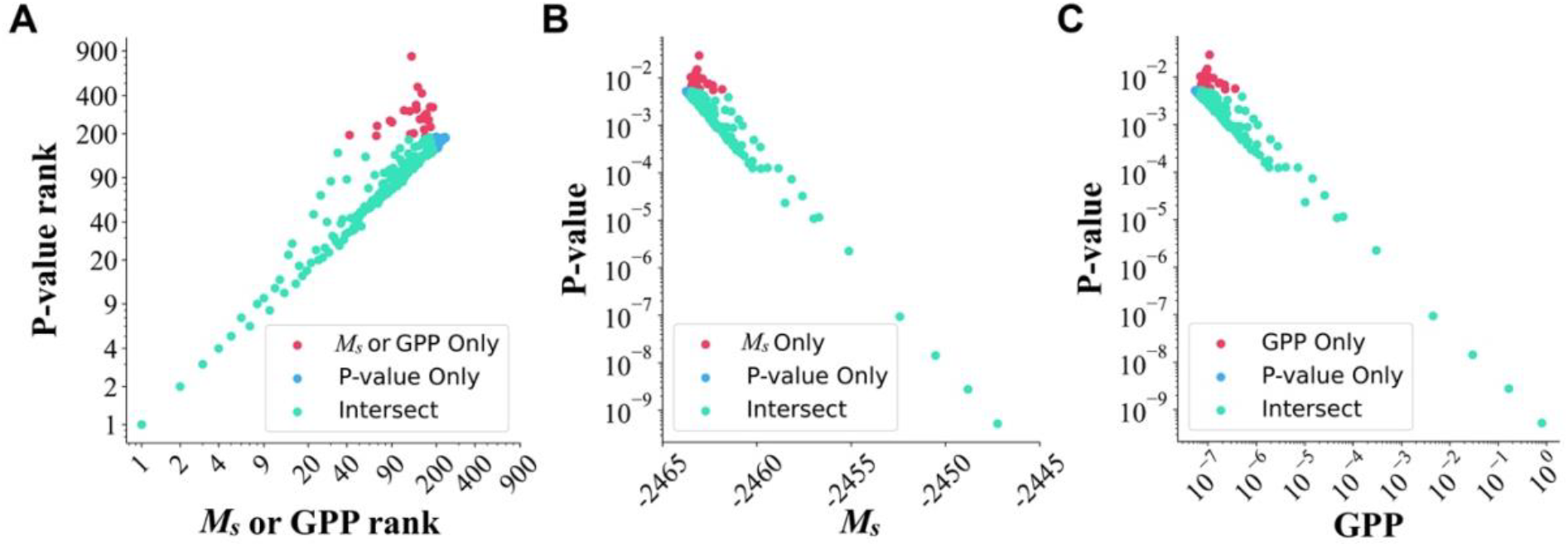
A Bayesian method for GWAS analysis. **A**. The p-value ranks and *M*_*s*_ or GPP ranks are the same or similar for the top 189 SNPs selected by *M*_*s*_ or p-value ranking. **B**. The p-values were very much negatively correlated with the *M*_*s*_ values of the top 189 SNPs selected by *M*_*s*_ or p-value ranking. **C**. The p-values were very much negatively correlated with the global posterior probabilities (GPP) of the top 189 SNPs selected by *M*_*s*_ or p-value ranking.

### IBI complements GWAS and better detects significant variants of low MAF

We applied both IBI and GWAS to the training (discovery) subset of 5,290 FHS subjects with 19,276 SNPs and HTN status, and derived the IBI marginal values of *M*_*s,r*_ (Figure 3A) and GWAS p-values (Figure 3B) for all the SNPs in the Manhattan plots. In Figure 3A, the values of *M*_*s,r*_ were normalized with (−2436 − M_*s*,r_) / (−2436 − (−2463)) considering -2436 as the maximum and -2463 as the minimum, based on the min-max normalization technique. For the GWAS analysis, if considering 0.05 / 19276 = 2.59e−6 as the significance level for p-value after the Bonferroni correction, five SNPs reach such significance (Figure 3B). In Figure 2, the population-level *M*_*s*_ values were derived by assuming one SNP as the global predictor or potential cause of HTN for the entire population (Figure 1B). When using two SNPs to specifically explain HTN status from two distinct subpopulations as is done by IBI (Figure 1C), the overall marginal values (*M*_*s,r*_) significantly increase for many SNPs of *V*_*s*_. Among all the SNPs, 189 *V*_*s*_ SNPs have *M*_*s,r*_ values bigger than the biggest *M*_*s*_ value derived in the population level from the best global predictor, represented as *V*_*g*_ (Figure 3A). The higher score of IBI compared to the population-based Bayesian method also has theoretical support. It has been proved that instance-based (i.e., individualized) causal inference methods, a family of algorithms to which the IBI belongs, are consistent. More specifically, in the large sample limit, the score of the data-generating instance-specific model will be assigned the highest score of any model [21]. These results support that the HTN status in the overall population has been explained better by IBI with any of the top 189 SNPs, *V*_*s*_, explaining the subpopulation of 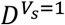 and with the remaining population predictor, *V*_*r*_, explaining the remaining 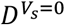 subpopulation, in comparison to using the best global predictor *V*_*g*_ itself to explain the entire population of *D*.

**Figure 3.**
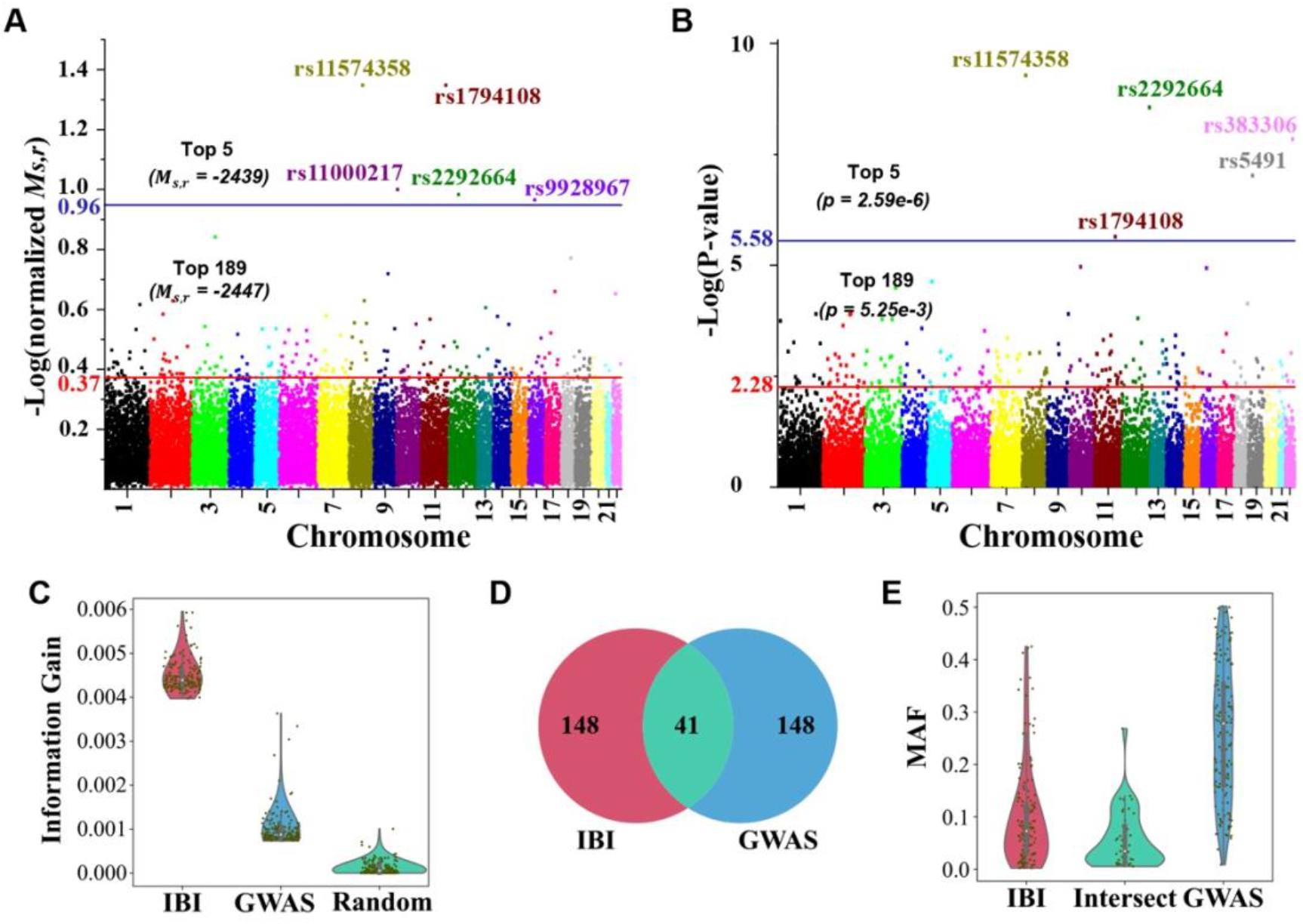
Comparison of IBI and GWAS. **A**. A Manhattan plot of SNPs’ chromosome location and normalized *M*_*s,r*_ values acquired by IBI. The threshold lines in blue and red together with the corresponding threshold *M*_*s,r*_ values were labeled for both top 5 and top 189 SNPs ranked by *M*_*s,r*_. Top five SNPs were annotated with rs IDs. **B**. A Manhattan plot of SNPs’ chromosome location and p-values acquired by GWAS. The threshold lines in blue and red together with the corresponding threshold p-values were labeled for both top 5 and top 189 SNPs ranked by p-values. Top five SNPs were annotated with rs IDs. **C**. Information gain from top 189 SNPs ranked by IBI, GWAS and randomly-selected 189 SNPs. The black dots represent the information gain values for individual SNPs. **D**. A Venn diagram of top 189 SNPs ranked by IBI and GWAS. **E**. Violin plots of the MAF distributions of the SNPs in the three sections of D, IBI, Intersection, GWAS. The black dots represent the MAF values for individual SNPs. The thick vertical gray bars show the interquartile range and the three white dots represent the medians. Wider sections suggest higher probability for the given MAF values.

We performed another evaluation from the perspective of information theory. In this setting, GWAS analysis is searching for a variant *V*_*s*_ that has strong information with respect to a trait *T*_*i*_ (HTN), and the amount of information can be measured as information gain (IG) [22]. IG can be calculated by splitting samples according to one variable (*V*_*s*_) and then measuring the change in the entropy of the other variable (*T*_*i*_) during partitions. Rather than focusing on just one variant as in a GWAS analysis, the IBI algorithm evaluates how much information we can gain with respect to the trait *T*_*i*_ (HTN) if we consider both the specific variant of interest (*V*_*s*_) and the remaining-population predictor (*V*_*r*_), IG (*V*_*s*_, *V*_*r*_ ; *T*_*i*_) [23]. The more *V*_*s*_ and *V*_*r*_ complement each other (e.g., they are two distinct predictors for different subgroups) to provide information with respect to *T*_*i*_, the higher the IG. In other words, IBI searches for variants that not only explain HTN well in the “patients-like-me” subpopulation, but also help enhance the information of *V*_*r*_ with respect to HTN in the remaining subpopulation that do not contain this specific variant of interest. Actually, the ranking of the top 189 SNPs by IBI *M*_*s,r*_ turned out to be highly correlated (Spearman correlation coefficient, r = 0.9) to their ranking by IG values: in general, the higher the *M*_*s,r*_, the higher the IG. Based on information gain values, top-5 IBI SNPs were rs11574358, rs1794108, rs11000217, rs2292664 and rs9928967 while top-5 GWAS SNPs were rs11574358, rs2292664, rs383306, rs5491 and rs1794108. IG values for the top 189 IBI SNPs of *V*_*s*_ selected by *M*_*s,r*_ are significantly higher than those of the top 189 GWAS SNPs selected by the p-values; IG values for the top 189 GWAS SNPs are also significantly higher than the values for 189 randomly-selected SNPs (Figure 3C) as expected.

We further examined whether there is any overlap between the top 189 IBI and GWAS SNPs. We found that 41 of the top 189 SNPs were identified by both IBI and GWAS (Figure 3D), and 3 of the top 5 IBI and GWAS SNPs are the same (Figure 3A, B). Thus, IBI and GWAS share many of the same top SNPs, suggesting a mutual agreement between these two approaches. The unique SNPs for IBI or GWAS support that the two approaches are also complementary (Fig. 3D). We further examined the MAF distribution for the different subsets in Figure 3D (Figure 3E). Interestingly, the IBI-only SNPs overall had much lower MAF than GWAS-only SNPs (Fig. 3E). This result provides support for the hypothesis that IBI is more capable of identifying lower-frequency significant variants, relative to GWAS, by concentrating on the genomic variants of a given individual in a specific subpopulation.

### IBI discovered more individualized and diverse significant SNPs that better explain the HTN patients, compared to GWAS

For a given individual *h*, IBI derives the posterior probability for each genomic variant 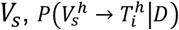, by normalizing *M*_*s,r*_ with a summation of all the *M*_*s,r*_ across all the existing minor allele SNPs (i.e., the SNP value is 1) in this individual. This posterior probability takes into consideration the diverse genomic background or context for different individuals. More specifically, a particular SNP *V*_*s*_ with the same *M*_*s,r*_ may have different posterior probabilities in different individuals due to their distinct genomic background (i.e., different sets of existing minor allele SNPs). For a given HTN patient, IBI ranked all the minor alleles existing in this individual based on their individualized posterior probabilities (this ranking will be the same as the ranking based on *M*_*s,r*_ in a given patient); the SNP with the highest posterior probability was considered as the most probable influence of HTN for this given patient. For comparison, we designated a top SNP for each HTN patient based on the population-level p-values derived by GWAS: among the existing minor alleles in a given HTN patient, the non-protective minor allele with the lowest (most significant) p-value was considered to be the most probable influence for HTN in this particular patient.

Among all the 930 HTN patients in the discovery dataset, we identified 16 unique SNPs according to GWAS ranking (Figure 4A) and 25 unique SNPs based on IBI ranking (Figure 4B); each of these unique SNPs was assigned by GWAS or IBI as a top-1 SNP for at least one HTN patient. The number of HTN patients explained by each of these unique SNP can be derived by the differences of the accumulated number of explained HTN patients showed in Figure 4A, B. The more unique SNPs identified by IBI suggested IBI was able to find a more diverse set of significant SNPs with a more personalized approach. IBI identified 13 SNPs that explain less than 10 HTN patients individually while GWAS found 6 such SNPs. Interestingly, at the same time, IBI assigns the intronic SNP ‘rs13265032’ in the *CSMD1* loci as the top-1 SNP with the highest PP or *M*_*s,r*_ for each of the 425 (46%) of HTN patients (Figure 4B).

**Figure 4.**
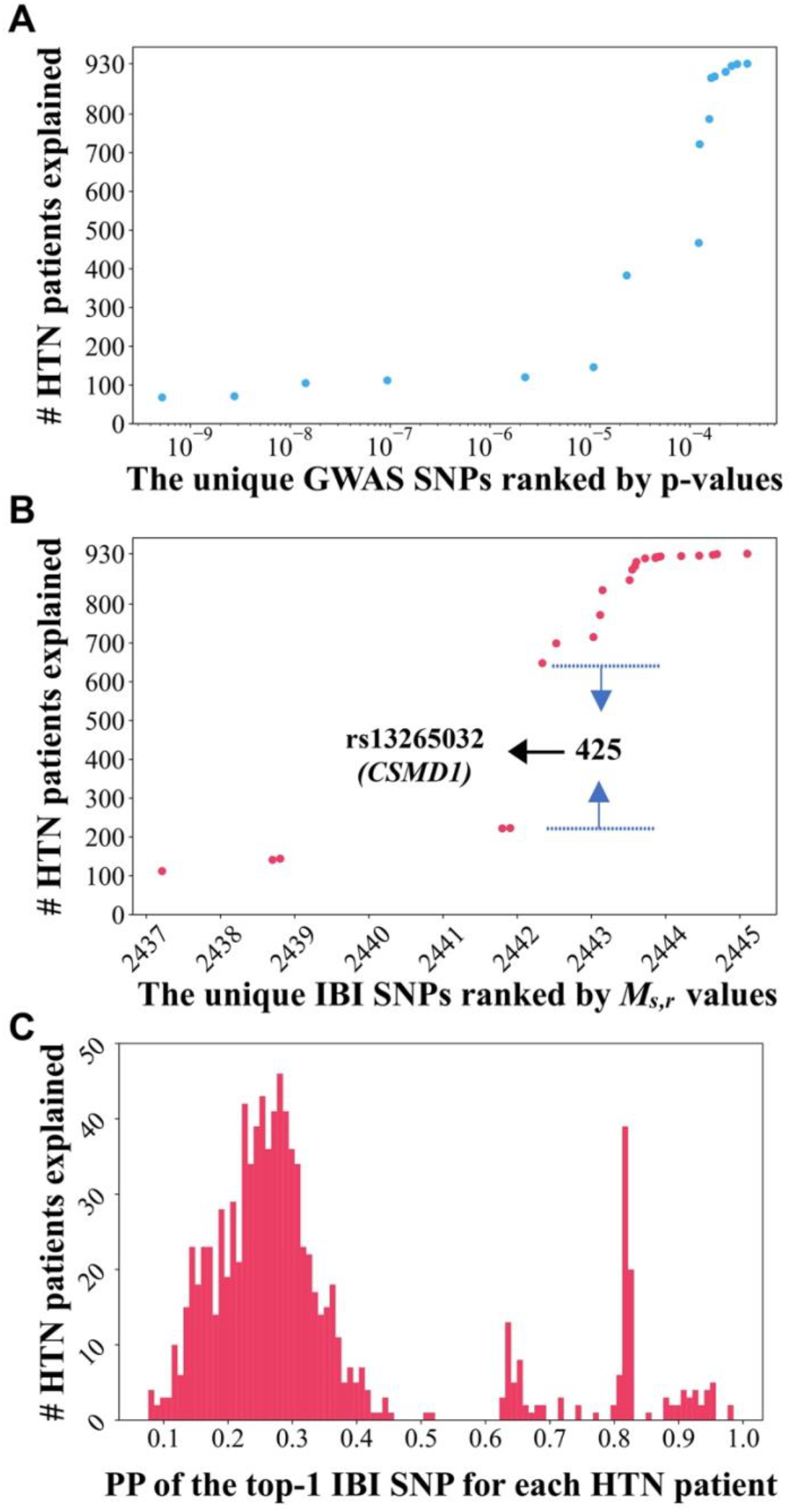
HTN patient coverage. **A**. The 16 unique GWAS SNPs ranked by p-values were plotted against the cumulative number of HTN patients explained. **B**. The 25 unique IBI SNPs ranked by *M*_*s,r*_ were plotted against the cumulative number of HTN patients explained. The number of HTN patients covered by each of these SNPs can be derived by taking the difference of the two adjacent cumulative numbers of explained HTN patients as showed for rs13265032 covering 425 HTN patients. **C**. Histogram of the individualized posterior probability of the top-1 SNP assigned by IBI to each of the 930 HTN patient.

For the GWAS analysis, if considering 0.05/19276 = 2.59e-6 as the significance level for p-value after the Bonferroni correction, then 120 out of 930 (12.9%) HTN patients can be assigned a significant SNP identified by GWAS; even with a relaxed significance level of 1.09e-5, only 146 out of 930 (15.7%) HTN patients are covered or explained by these significant GWAS SNPs (Figure 4A). This suggests that the significant SNPs of HTN identified at the population level by GWAS do not necessarily exist in a given HTN patient, leaving a significant portion of HTN patients unexplained by these significant SNPs.

On average, there are 7,767 out of 19,276 minor allele SNPs existing in the HTN patients. If assuming all these existing risk alleles have the same prior probability in causing HTN, and only one of them is causing HTN, then the prior probability for each risk allele is 1.0 / 7767 = 1.3e-4. Interestingly, the top one SNP selected by IBI for each HTN patient has much higher posterior probability ranging from 0.08 to 0.99 (Figure 4C). If considering 0.1 as a significant posterior probability threshold, then 922 out of 930 (99.1%) HTN patients can be assigned a significant IBI SNP as the potential cause for their HTN status; with a more restrictive threshold of 0.2, 741 out of 930 (79.7%) HTN patients can be explained. These results suggested that IBI was able to find a top SNP with significant posterior probability (>=0.1), relative to the random chance (1.3e-4), for the majority of HTN patients as a potential genomic cause.

As is shown in Table 1, the intronic SNP ‘rs13265032’ in the *CSMD1* loci that was assigned as the top-1 SNP by IBI for 46% (425) of HTN patients, was also ranked high (12) by IBI *M*_*s,r*_ among all the SNPs. By contrast, this SNP was never assigned as a top-1 SNP for any HTN patient by GWAS and this SNP was ranked very low (5361) by GWAS among all the SNPs. Interestingly, other intronic SNPs in the *CSMD1* loci have been reported to be associated with hypertension [24] or blood pressure response to hydrochlorothiazide [25, 26], an antihypertensive drug. Among all the 86 SNPs located in the *CSMD1* loci in our dataset, three of them were ranked very high by IBI as is shown in Table 1, while all of them were ranked relatively low by GWAS. These three novel SNPs identified by IBI in the *CSMD1* loci provide evidence to support the reported role of *CSMD1* in HTN, which themselves may warrant further analysis for their potential causal influence on *CSMD1* regulation. In addition to SNPs in the *CSMD1* loci, IBI also identified a novel missense variant of ‘rs1803274’ in the *BCHE* loci, a novel intron variant of ‘rs948028’ in the *GRIK4* loci and a novel missense variant of ‘rs12779623’ in the *MALRD1* loci as top-1 likely cause of HTN in 9, 2 and 1 HTN patient, respectively (Table 1). Interestingly, *BCHE* [27, 28] loci, *GRIK4* [29] loci and *MALRD1* loci [4] have been reported to be associated with blood pressure regulation, although GWAS analysis ranked their SNPs relatively low (Table 1). Overall, these results provide support for IBI being able to identify novel and biologically meaningful SNPs or genes associated with HTN that were missed by GWAS analysis.

**Table 1.**
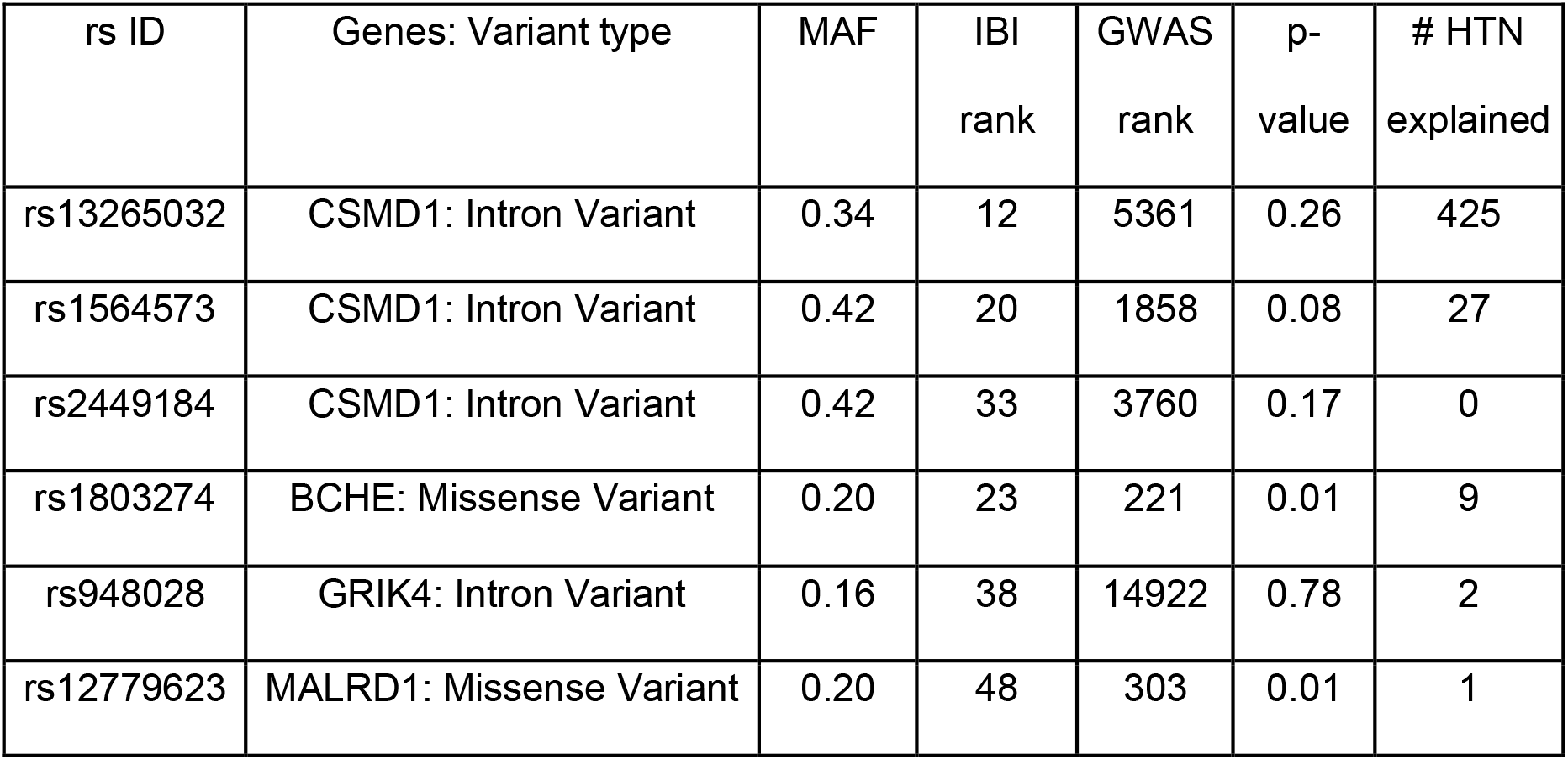
Novel SNPs in BP-associated genes identified by IBI as the individualized and most-probable HTN cause.

### IBI found well-known significant variants or genes that were missed by the parallel GWAS analysis in the same cohort

We list several missense variants (Table 2) as well as the gene loci (Table 3) that were previously reported for their influence on blood pressure regulation, in addition to the ones discussed in Table 1. In Tables 2 and 3, IBI ranks were determined by *M*_*s,r*_ while GWAS ranks were determined by the p-value.

**Table 2.**
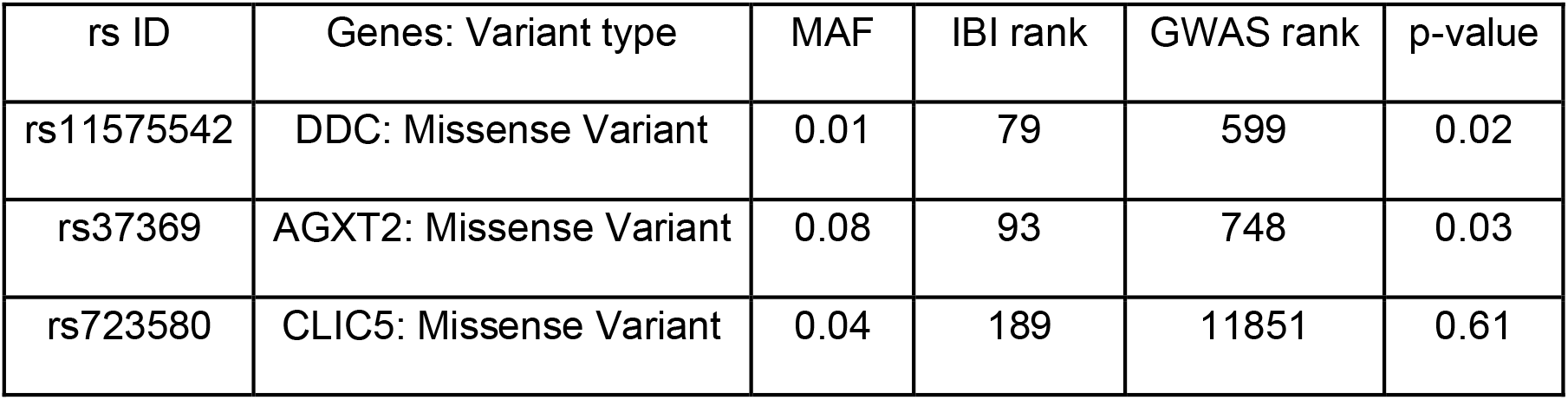
SNPs well-known for blood pressure regulation identified by IBI but missed by GWAS.

| rs ID | Genes: Variant type | MAF | IBI rank | GWAS rank | p-value |
| --- | --- | --- | --- | --- | --- |
| rs11575542 | DDC: Missense Variant | 0.01 | 79 | 599 | 0.02 |
| rs37369 | AGXT2: Missense Variant | 0.08 | 93 | 748 | 0.03 |
| rs723580 | CLIC5: Missense Variant | 0.04 | 189 | 11851 | 0.61 |

**Table 3.** Genes well-known for blood pressure regulation identified by IBI but ranked relatively low by GWAS.

| rs ID | Genes: Variant type | MAF | IBI rank | GWAS rank | Top GWAS rank | p-value |
| --- | --- | --- | --- | --- | --- | --- |
| rs3211938 | CD36 [36, 37]: Stop Gained | 0.01 | 32 | 141 | 141 | 3.8E-03 |
| rs6730396 | ALLC [38]: Missense Variant | 0.01 | 45 | 192 | 192 | 5.5E-03 |
| rs9896904 | ANKFN1 [39]: Intron Variant | 0.07 | 57 | 14392 | 2130 | 7.5E-01 |
| rs11899922 | THSD7B [4, 40]: Intron Variant | 0.07 | 70 | 9415 | 929 | 4.8E-01 |
| rs10968668 | LINGO2 [41, 42]: Intron Variant | 0.08 | 80 | 1237 | 1237 | 4.9E-02 |
| rs13261739 | PDGFRL [43, 44]: Intron Variant | 0.13 | 94 | 7156 | 7156 | 3.6E-01 |
| rs6140644 | PLCB1 [45]: Intron Variant | 0.17 | 116 | 9947 | 699 | 5.1E-01 |
| rs7647302 | KCNAB1 [46]: Intron Variant | 0.01 | 158 | 9528 | 1964 | 4.9E-01 |

In Table 2, the missense variant of rs37369 [30] has been shown to be one of the 4 functional SNPs of AGXT2, which has been reported to have strong associations with several cardiorenal traits, such as coronary heart disease [31]. Its significant association with hypertension was very recently reported via multiple regression analysis involving only several targeted SNPs [32]. The missense variant rs11575542 was very recently identified as a functional variant of the DOPA Decarboxylase (*DDC*) gene during the systematic polymorphism screening across the 15-Exon *DCC* locus [33]. The SNP was shown to alter the enzyme activity of DCC and result in changes in renal DA excretion that is linked to hypertension [33]. The missense variant of rs723580 was reported to be a top trans-eSNP [34] for the expression level of EPO associated with the red blood cell traits that were strongly linked to hypertension [35]. Intriguingly, with low MAF in our relatively small discovery cohort, these three SNPs were ranked much higher by IBI than by the parallel GWAS analysis (Table 2). This result provides support that IBI can recognize biologically meaningful genomic variants of low MAF, relative to GWAS, particularly when the sample size is small compared to the number of SNPs to be tested.

Table 3 shows a list of genes that have been reported to be associated with BP regulation or HTN where at least one related paper is listed for each gene. IBI identified these genes as candidate genes influencing HTN since these gene loci contain at least one novel SNP that is highly ranked by IBI for its association with HTN. Interestingly, all of these genes were ranked relatively low by GWAS even considering the highest SNP rank by GWAS (‘Top GWAS rank’ in Table 3) within each gene locus; all of these novel SNPs were also ranked relatively low by GWAS (‘GWAS rank’ in Table 3) with non-significant p-values (Table 3). Moreover, among these eight SNPs highly-ranked by IBI and lowly-ranked by GWAS, six have MAF lower than 0.1 and three have MAF as low as 0.01 (Table 3). This together with Table 2 supports that IBI is more capable in identifying significant variants of low MAF, compared to GWAS.

### IBI top SNPs identify genetic risk scores that are more predictive for HTN than do the GWAS top SNPs

We further compared the capabilities of significant SNPs, identified by IBI and GWAS, in predicting HTN. After running IBI and GWAS on the training (discovery) dataset, we were able to rank all the SNPs based on *M*_*s,r*_ derived from IBI or p-values obtained from GWAS. For each subject in the test set, based on IBI ranking or GWAS ranking, we identified the top 1 and 3 SNPs that exist in this subject (with a value of ‘1’ denoting a minor allele). We then used these top SNPs to calculate the genetic risk scores (GRS) for each subject by the sum of his or her risk alleles, weighted by odds ratio for GWAS top SNPs or by *M*_*s,r*_ for IBI top SNPs. We further use min-max normalization to normalize both the IBI and GWAS GRS to avoid potential bias from the different scales of the original values. We then directly calculated the area under ROC curve (AUROC or AUC) using the normalized GRS for each patient (Figure 5). We also trained a logistic regression model for HTN prediction using this feature of normalized GRS, which gave very similar results (data not shown).

**Figure 5.**
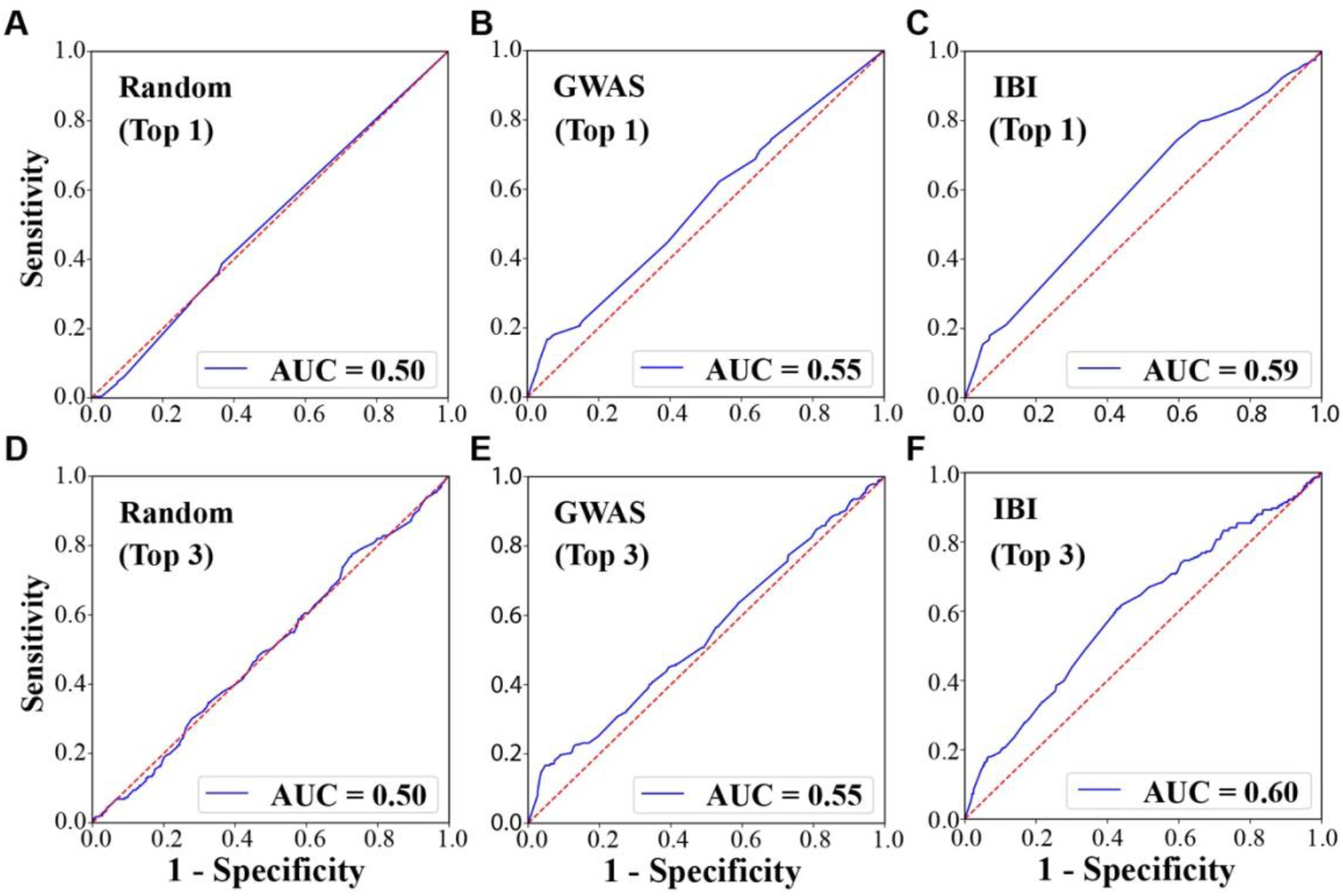
ROC curves for predicting hypertension from top SNPs. Top 1 SNP (A, B, C) and top 3 SNPs (D, E, F) selected by GWAS (p-value ranking), IBI (*M*_*s,r*_ ranking) or randomly were used to calculate the genetic risk scores for each test patient, which were further used to derive the AUROC for predicting hypertension. For these two experiments, random SNPs have AUROC as 0.50 (Top 1/3 SNP; (A, D); GWAS-selected top SNPs have AUROC as 0.55 (B, E); IBI-selected top SNPs have AUROC as 0.59 for top 1 SNP and 0.60 for top 3 SNPs.

As expected, using randomly selected SNPs showed poor prediction performance, with an AUROC of 0.50 (Figure 5A, D); the GWAS-selected top one or three SNPs both have an AUROC of 0.55 (Figure 5B, E) suggesting some level of prediction; the IBI-selected top SNP had an AUROC of 0.59 (Figure 5C) and the top 3 SNPs had an AUROC of 0.60 (Figure 5F). The higher AUROC achieved by IBI provides support that the top SNPs it selects predict hypertension better than the top SNPs selected by GWAS.

## Discussion

In this study, we developed and applied a novel and individualized method (IBI) to estimate the personalized genomic variants for the complex trait of hypertension. We compared its performance with the population-based GWAS method using a real dataset from the FHS cohort. The significant overlap of the top-ranked SNPs by both IBI and GWAS suggest a degree of agreement of these two approaches. On the other hand, the unique SNPs they found support a complementary role of IBI to current GWAS analyses. Interestingly, by focusing on each individual and its patient-like-me subgroup, IBI was capable of identifying significant SNPs of low MAF in the same cohort, relative to GWAS. IBI was also able to identify more diverse and individualized top SNPs to explain the HTN patients. Moreover, the top SNPs identified by IBI from the discovery cohort were able to predict HTN better than the top ones derived from GWAS when applied to an unseen test cohort. We also identified evidence from the literature to support the biological significance of top SNPs found by IBI, especially the ones highly-ranked by IBI and lowly-ranked by GWAS. In summary, our study provides support that IBI can serve as a complementary approach in discovering novel and personalized genomic variants that may be missed by GWAS.

Contemporary GWAS studies often involve a large sample size (∼1 million) to gain sufficient power, especially for variants of low MAF. Considering the large genomic heterogeneity among different individuals, as well as the nature of complex diseases often being affected by many variants of small effect size, an alternative approach is to focus on the subpopulation containing the specific variant of low MAF under evaluation, as IBI does. In this way, IBI may be able to better evaluate the effect of low-MAF variants in a patient-like-me subpopulation, without the potential noise from a large remaining population not containing such variants; moreover, this large remaining population could be explained better with a remaining population driver. The fact that the top-ranked SNPs by IBI in general have a higher overall marginal likelihood, *M*_*s,r*_, and higher information gain with respect to the HTN status, provide support that IBI may have found specific drivers that better explain the subpopulations. Our results also support that IBI is not compromised in identifying significant high-MAF SNPs. IBI’s population partition strategy aligns well with the concept of personalized medicine in which different individuals or subpopulations may have different underlying genomic influences on producing complex clinical phenotypes such as HTN.

As a general Bayesian framework, IBI can be applied to any discrete trait. It can also be applied to continuous traits by changing the marginal likelihood function from using the BDeu score for discrete variables to using the Bayesian information criterion (BIC) score for continuous variables [14]. For the current approach presented in this study, one limitation is that it only considers the genomic factors of HTN, while not modeling the effects of other factors such as age, sex, population structure and the family relatedness that may exist in this FHS cohort. To model the effects from non-genomic factors, we plan to incorporate linear mixed models [47-51] into our current framework. Also, due to confounding factors such as population structure, as well as linage disequilibrium (LD), the predictive variants described in this paper are not guaranteed to be causal. Further fine mapping approaches, functional analysis, or Mendelian randomization can be used to further pinpoint the potential causality. Another interesting future direction is to search for more than one genomic variant that might work together to affect and predict the phenotypes of individuals and subpopulations.

## Conclusions

In summary, we described a novel Bayesian method for identifying personalized genomic variants that predict complex traits, such as HTN. IBI can serve as a complementary approach to GWAS, especially in detecting significant genomic variants of low frequency. The novel SNPs we identified for HTN warrant further analysis for their possible causal role in blood pressure regulation.

## Data Availability

All data produced in the present study are available upon reasonable request to the authors

https://www.ncbi.nlm.nih.gov/projects/gap/cgi-bin/study.cgi?study_id=phs000007.v32.p13

## List of abbreviations

GWAS: Genome Wide Association Study
IBI: Individualized Bayesian inference
TCI: tumor-specific causal inference
HTN: Hypertension
BP: Blood pressure
SBP: Systolic blood pressure
DBP: Diastolic blood pressure
FHS: Framingham Heart Study
MAF: Minor allele frequency
CBN: Causal Bayesian network
BDeu: Bayesian Dirichlet equivalent uniform
TOPMed: The Trans-Omics for Precision Medicine
IG: Information gain
GRS: Genetic risk score
AUROC or AUC: area under ROC curve
BIC: Bayesian information criterion

## Declarations

### Ethics approval and consent to participate

Not applicable.

### Consent for publication

Not applicable.

### Availability of data and materials

The genomic and blood pressure data from the FHS cohort (study accession: phs000007.v30.p11) are publicly available via controlled-access through the database of Genotypes and Phenotypes Study (https://www.ncbi.nlm.nih.gov/projects/gap/cgi-bin/study.cgi?study_id=phs000007.v30.p11). The source code for IBI is publicly available on GitHub at https://github.com/asadcfc/IBI.

### Authors’ contributions

JL designed and implemented the IBI algorithm in python, conceived and designed the experiments, and wrote the paper; MA.R and JL carried out the data collection, modeling, analyses; XL and GC provided insightful advices about the design of the IBI algorithm and some of the experiments, as well as edited the manuscript; YD provided advice for whole-genome data processing and GWAS analysis; DM provided general advice on hypertension development and its potential genomic causes; CC helped with the information gain experiment; All authors read and approved the final manuscript.

## Acknowledgements

The Framingham Heart Study, within the database of Genotypes and Phenotypes Study (accession #: phs000007.v30.p11), is conducted and supported by the National Heart, Lung, and Blood Institute (NHLBI) in collaboration with Boston University (Contract No. N01-HC-25195 and HHSN268201500001I). This manuscript was not prepared in collaboration with investigators of the Framingham Heart Study and does not necessarily reflect the opinions or views of the Framingham Heart Study, Boston University, or NHLBI. Funding for SHARe Affymetrix genotyping was provided by NHLBI Contract N02-HL-64278. SHARe Illumina genotyping was provided under an agreement between Illumina and Boston University. Funding for Affymetrix genotyping of the FHS Omni cohorts was provided by Intramural NHLBI funds from Andrew D. Johnson and Christopher J. O’Donnell.

Support for this work was provided by the National Institutes of Health, National Heart, Lung, and Blood Institute, through the BioData Catalyst program (award 1OT3HL142479-01, 1OT3HL142478-01, 1OT3HL142481-01, 1OT3HL142480-01, 1OT3HL147154-01). Any opinions expressed in this document are those of the author(s) and do not necessarily reflect the views of NHLBI, individual BioData Catalyst team members, or affiliated organizations and institutions. The authors wish to acknowledge the contributions of the consortium working on the development of the NHLBI BioData Catalyst ecosystem.

## References

1. Padmanabhan S, Joe B: Towards Precision Medicine for Hypertension: A Review of Genomic, Epigenomic, and Microbiomic Effects on Blood Pressure in Experimental Rat Models and Humans. Physiol Rev 2017, 97(4):1469–1528.

2. Padmanabhan S, Dominiczak AF: Genomics of hypertension: the road to precision medicine. Nat Rev Cardiol 2021, 18(4):235–250.

3. Cabrera CP, Ng FL, Nicholls HL, Gupta A, Barnes MR, Munroe PB, Caulfield MJ: Over 1000 genetic loci influencing blood pressure with multiple systems and tissues implicated. Hum Mol Genet 2019, 28(R2):R151-R161.

4. Evangelou E, Warren HR, Mosen-Ansorena D, Mifsud B, Pazoki R, Gao H, Ntritsos G, Dimou N, Cabrera CP, Karaman I et al: Genetic analysis of over 1 million people identifies 535 new loci associated with blood pressure traits. Nat Genet 2018, 50(10):1412–1425.

5. Surendran P, Feofanova EV, Lahrouchi N, Ntalla I, Karthikeyan S, Cook J, Chen L, Mifsud B, Yao C, Kraja AT et al: Discovery of rare variants associated with blood pressure regulation through meta-analysis of 1.3 million individuals. Nat Genet 2020, 52(12):1314–1332.

6. Giri A, Hellwege JN, Keaton JM, Park J, Qiu C, Warren HR, Torstenson ES, Kovesdy CP, Sun YV, Wilson OD et al: Trans-ethnic association study of blood pressure determinants in over 750,000 individuals. Nat Genet 2019, 51(1):51–62.

7. Russo A, Di Gaetano C, Cugliari G, Matullo G: Advances in the Genetics of Hypertension: The Effect of Rare Variants. Int J Mol Sci 2018, 19(3).

8. Banerjee S, Zeng L, Schunkert H, Soding J: Bayesian multiple logistic regression for case-control GWAS. PLoS Genet 2018, 14(12):e1007856.

9. Schaid DJ, Chen W, Larson NB: From genome-wide associations to candidate causal variants by statistical fine-mapping. Nat Rev Genet 2018, 19(8):491–504.

10. Stephens M, Balding DJ: Bayesian statistical methods for genetic association studies. Nat Rev Genet 2009, 10(10):681–690.

11. Cai C, Cooper GF, Lu KN, Ma X, Xu S, Zhao Z, Chen X, Xue Y, Lee AV, Clark N et al: Systematic discovery of the functional impact of somatic genome alterations in individual tumors through tumor-specific causal inference. PLoS Comput Biol 2019, 15(7):e1007088.

12. Heckerman D, Meek C, Cooper G: A Bayesian approach to causal discovery. In: Computation, causation, and discovery (MIT Press) Edited by Glymour C, Cooper G, vol. 19; 1999: 141–166.

13. Spirtes P GC, Scheines R.: Causation, Prediction, and Search.: Cambridge, MA: MIT Press;; 2000.

14. Heckerman D, Geiger D, Chickering DM: Learning Bayesian networks: The combination of knowledge and statistical data. Machine learning 1995, 20(3):197–243.

15. Consortium BC: The NHLBI BioData Catalyst. Zenodo(31 August 2020, date last accessed) 2020.

16. Gogarten SM, Sofer T, Chen H, Yu C, Brody JA, Thornton TA, Rice KM, Conomos MP: Genetic association testing using the GENESIS R/Bioconductor package. Bioinformatics 2019, 35(24):5346–5348.

17. Mahmood SS, Levy D, Vasan RS, Wang TJ: The Framingham Heart Study and the epidemiology of cardiovascular disease: a historical perspective. The Lancet 2014, 383(9921):999–1008.

18. Tsao CW, Vasan RS: Cohort Profile: The Framingham Heart Study (FHS): overview of milestones in cardiovascular epidemiology. Int J Epidemiol 2015, 44(6):1800–1813.

19. Stilp AM, Emery LS, Broome JG, Buth EJ, Khan AT, Laurie CA, Wang FF, Wong Q, Chen D, D’Augustine CM et al: A System for Phenotype Harmonization in the National Heart, Lung, and Blood Institute Trans-Omics for Precision Medicine (TOPMed) Program. Am J Epidemiol 2021, 190(10):1977–1992.

20. Levy D, Ehret GB, Rice K, Verwoert GC, Launer LJ, Dehghan A, Glazer NL, Morrison AC, Johnson AD, Aspelund T et al: Genome-wide association study of blood pressure and hypertension. Nat Genet 2009, 41(6):677–687.

21. Jabbari F: Instance-Specific Causal Bayesian Network Structure Learning. University of Pittsburgh; 2021.

22. Kent JT: Information gain and a general measure of correlation. Biometrika 1983, 70(1):163–173.

23. Quinlan JR: Induction of decision trees. Machine learning 1986, 1(1):81–106.

24. Hong KW, Go MJ, Jin HS, Lim JE, Lee JY, Han BG, Hwang SY, Lee SH, Park HK, Cho YS et al: Genetic variations in ATP2B1, CSK, ARSG and CSMD1 loci are related to blood pressure and/or hypertension in two Korean cohorts. Journal of Human Hypertension 2010, 24(6):367–372.

25. Chittani M, Zaninello R, Lanzani C, Frau F, Ortu MF, Salvi E, Fresu G, Citterio L, Braga D, Piras DA et al: TET2 and CSMD1 genes affect SBP response to hydrochlorothiazide in never-treated essential hypertensives. J Hypertens 2015, 33(6):1301–1309.

26. Salvi E, Wang Z, Rizzi F, Gong Y, McDonough CW, Padmanabhan S, Hiltunen TP, Lanzani C, Zaninello R, Chittani M et al: Genome-Wide and Gene-Based Meta-Analyses Identify Novel Loci Influencing Blood Pressure Response to Hydrochlorothiazide. Hypertension 2017, 69(1):51–59.

27. Benyamin B, Middelberg RP, Lind PA, Valle AM, Gordon S, Nyholt DR, Medland SE, Henders AK, Heath AC, Madden PAF et al: GWAS of butyrylcholinesterase activity identifies four novel loci, independent effects within BCHE and secondary associations with metabolic risk factors. Human Molecular Genetics 2011, 20(22):4504–4514.

28. Cardoso AM, Abdalla FH, Bagatini MD, Martins CC, Fiorin Fda S, Baldissarelli J, Costa P, Mello FF, Fiorenza AM, Serres JD et al: Swimming training prevents alterations in acetylcholinesterase and butyrylcholinesterase activities in hypertensive rats. Am J Hypertens 2014, 27(4):522–529.

29. Rutherford S, Cai G, Lopez-Alvarenga JC, Kent JW, Voruganti VS, Proffitt JM, Curran JE, Johnson MP, Dyer TD, Jowett JB: A chromosome 11q quantitative-trait locus influences change of blood-pressure measurements over time in Mexican Americans of the San Antonio Family Heart Study. The American Journal of Human Genetics 2007, 81(4):744–755.

30. Suhre K, Wallaschofski H, Raffler J, Friedrich N, Haring R, Michael K, Wasner C, Krebs A, Kronenberg F, Chang D: A genome-wide association study of metabolic traits in human urine. Nature genetics 2011, 43(6):565–569.

31. Hu XL, Zeng WJ, Li MP, Yang YL, Kuang DB, Li H, Zhang YJ, Jiang C, Peng LM, Qi H et al: AGXT2 rs37369 polymorphism predicts the renal function in patients with chronic heart failure. Gene 2017, 637:145–151.

32. Yoshino Y, Kumon H, Mori T, Yoshida T, Tachibana A, Shimizu H, Iga J-i, Ueno S-i: Effects of AGXT2 variants on blood pressure and blood sugar among 750 older Japanese subjects recruited by the complete enumeration survey method. BMC Genomics 2021, 22(1):287.

33. Miramontes-Gonzalez JP, Hightower CM, Zhang K, Kurosaki H, Schork AJ, Biswas N, Vaingankar S, Mahata M, Lipkowitz MS, Nievergelt CM et al: A new common functional coding variant at the DDC gene change renal enzyme activity and modify renal dopamine function. Scientific Reports 2019, 9(1):5055.

34. Zhang X, Johnson AD, Hendricks AE, Hwang S-J, Tanriverdi K, Ganesh SK, Smith NL, Peyser PA, Freedman JE, O’Donnell CJ: Genetic associations with expression for genes implicated in GWAS studies for atherosclerotic cardiovascular disease and blood phenotypes. Human Molecular Genetics 2013, 23(3):782–795.

35. Tsuda K: Red blood cell abnormalities and hypertension. Hypertension Research 2020, 43(1):72–73.

36. Momeni-Moghaddam MA, Asadikaram G, Akbari H, Abolhassani M, Masoumi M, Nadimy Z, Khaksari M: CD36 gene polymorphism rs1761667 (G> A) is associated with hypertension and coronary artery disease in an Iranian population. BMC cardiovascular disorders 2019, 19(1):1–9.

37. Pravenec M, Churchill PC, Churchill MC, Viklicky O, Kazdova L, Aitman TJ, Petretto E, Hubner N, Wallace CA, Zimdahl H: Identification of renal Cd36 as a determinant of blood pressure and risk for hypertension. Nature genetics 2008, 40(8):952–954.

38. Sung YJ, Basson J, Cheng N, Nguyen K-DH, Nandakumar P, Hunt SC, Arnett DK, Dávila-Román VG, Rao DC, Chakravarti A: The role of rare variants in systolic blood pressure: analysis of ExomeChip data in HyperGEN African Americans. Human heredity 2015, 79(1):20–27.

39. Chittani M, Zaninello R, Lanzani C, Frau F, Ortu MF, Salvi E, Fresu G, Citterio L, Braga D, Piras DA: TET2 and CSMD1genes affect SBP response to hydrochlorothiazide in never-treated essential hypertensives. Journal of hypertension 2015, 33(6):1301.

40. de Las Fuentes L, Sung YJ, Schwander KL, Kalathiveetil S, Hunt SC, Arnett DK, Rao D: The role of SNP-loop diuretic interactions in hypertension across ethnic groups in HyperGEN. Frontiers in genetics 2013, 4:304.

41. Parmar PG, Taal HR, Timpson NJ, Thiering E, Lehtimäki T, Marinelli M, Lind PA, Howe LD, Verwoert G, Aalto V: International genome-wide association study consortium identifies novel loci associated with blood pressure in children and adolescents. Circulation: Cardiovascular Genetics 2016, 9(3):266–278.

42. Feitosa MF, Kraja AT, Chasman DI, Sung YJ, Winkler TW, Ntalla I, Guo X, Franceschini N, Cheng C-Y, Sim X: Novel genetic associations for blood pressure identified via gene-alcohol interaction in up to 570K individuals across multiple ancestries. PloS one 2018, 13(6):e0198166.

43. Yang W, Huang J, Yao C, Fan Z, Ge D, Gan W, Huang G, Hui R, Shen Y, Qiang B: Evidence for linkage and association of the markers near the LPL gene with hypertension in Chinese families. Journal of medical genetics 2003, 40(5):e57–e57.

44. Chun HJ, Bonnet S, Chan SY: Translational advances in the field of pulmonary hypertension. Translating MicroRNA biology in pulmonary hypertension. It will take more than “miR” words. American journal of respiratory and critical care medicine 2017, 195(2):167–178.

45. Warren HR, Evangelou E, Cabrera CP, Gao H, Ren M, Mifsud B, Ntalla I, Surendran P, Liu C, Cook JP: Genome-wide association analysis identifies novel blood pressure loci and offers biological insights into cardiovascular risk. Nature genetics 2017, 49(3):403–415.

46. McCarthy NS, Vangjeli C, Cavalleri GL, Delanty N, Shianna KV, Surendran P, O’Brien E, Munroe PB, Masca N, Tomaszewski M: Two further blood pressure loci identified in ion channel genes with a genecentric approach. Circulation: Cardiovascular Genetics 2014, 7(6):873–879.

47. Chen H, Wang C, Conomos MP, Stilp AM, Li Z, Sofer T, Szpiro AA, Chen W, Brehm JM, Celedon JC et al: Control for population structure and relatedness for binary traits in genetic association studies via logistic mixed models. Am J Hum Genet 2016, 98(4):653–666.

48. Lippert C, Listgarten J, Liu Y, Kadie CM, Davidson RI, Heckerman D: FaST linear mixed models for genome-wide association studies. Nat Methods 2011, 8(10):833–835.

49. Pirinen M, Donnelly P, Spencer CCA: Efficient computation with a linear mixed model on large-scale data sets with applications to genetic studies. The Annals of Applied Statistics 2013, 7(1).

50. Wen YJ, Zhang H, Ni YL, Huang B, Zhang J, Feng JY, Wang SB, Dunwell JM, Zhang YM, Wu R: Methodological implementation of mixed linear models in multi-locus genome-wide association studies. Brief Bioinform 2018, 19(4):700–712.

51. Yu J, Pressoir G, Briggs WH, Vroh Bi I, Yamasaki M, Doebley JF, McMullen MD, Gaut BS, Nielsen DM, Holland JB et al: A unified mixed-model method for association mapping that accounts for multiple levels of relatedness. Nat Genet 2006, 38(2):203–208.

